# A Novel Box for Aerosol and Droplet Guarding and Evacuation in Respiratory Infection (BADGER) for COVID-19 and Future Outbreaks

**DOI:** 10.1101/2020.05.09.20096032

**Authors:** Hau D. Le, Gordon A. Novak, Kevin C. Janek, Jesse Wang, Khang N. Huynh, Chris Myer, Adam Weinstein, Erick L. Oberstar, Jim Rasmussen, Timothy H. Bertram

**Affiliations:** Department of Surgery, University of Wisconsin School of Medicine and Public Health, Madison, WI; Department of Pediatrics, University of Wisconsin School of Medicine and Public Health, Madison, WI; Department of Biomedical Engineering, University of Wisconsin – Madison, WI; Department of Chemistry, University of Wisconsin – Madison, WI; University of Wisconsin School of Medicine and Public Health; Sector 67, Madison, Wisconsin; Department of Mechanical Engineering, University of Wisconsin – Madison, WI; Department of Anesthesia, University of Wisconsin School of Medicine and Public Health, Madison, WI; Department of Civil and Environmental Engineering, University of Wisconsin – Madison, WI

## Abstract

The coronavirus disease 2019 (COVID-19) pandemic caused by the severe acute respiratory syndrome coronavirus-2 (SARS-CoV-2) has infected millions and killed hundreds of thousands of people worldwide as of May 2020. Healthcare providers are at increased risks of infection when caring for patients with COVID-19. The mechanism of transmission of SARS-CoV-2 is still not fully understood. However, there is growing evidence for airborne spread, in addition to direct droplet and indirect contact. Here, we report on the design, construction, and testing of the BADGER (Box for Aerosol and Droplet Guarding and Evacuation in Respiratory Infection), an affordable, scalable device that contains droplets and aerosol particles. A semi-sealed environment is created inside the BADGER, which maintains at least twelve-air changes per hour using in-wall vacuum suction, and multiple hand-ports enable healthcare providers to perform essential tasks on a patient’s airway and head. Overall, the BADGER has the potential to contain large droplets and small airborne particles as demonstrated by simulated qualitative and quantitative assessments to provide an additional layer of protection for healthcare providers treating COVID-19 patients.

## Introduction

The coronavirus disease 2019 (COVID-19) pandemic caused by the severe acute respiratory syndrome coronavirus-2 (SARS-CoV-2) has infected more than 3.5 million people worldwide and caused more than 250,000 deaths by early May 2020.^1^ In the battle against the pandemic, healthcare professionals and workers are particularly vulnerable to infection due to their close proximity to patients with COVID-19. As critical players in this pandemic, the safety of health care providers is essential for the survival of our global health systems. Current recommended infection control precautions for health care professionals include the use of personal protective equipment (PPE) such as N95 respirator masks, gloves, isolation gowns, face shields or goggles, and meticulous hand hygiene.^2^ Despite precautions, many health care professionals have become infected. Data from Italy indicates approximately 20% of Italian health care professionals have become infected with many succumbing to COVID-19.^3^ In the U.S, as of 4/9/2020, there are 9,282 health care workers who have COVID-19.^4^ In addition, expansion of the COVID-19 pandemic could lead to more instances of overwhelmed hospital environments and shortages of PPE, which would likely further increase the risks to health care workers.

As the infections multiply, definitive knowledge about how SARS-CoV-2 spreads remains scant.^5^ In general, it is thought that viral respiratory infections such as COVID-19 are spread by direct and indirect droplet contact.^6^ However, there is growing evidence that the SARS-CoV-2 virus can also be spread by airborne transmission, similar to SARS-CoV-1 (**Figure 1A**).^7–11^ The size of the infective virion is believed to be approximately 80-160 nm, similar to SARS-CoV-1 and other coronaviruses. Most particles generated during coughing and sneezing are between 0.35 μm and 10 μm.^12^ Direct droplet spread occurs when droplets from a patient’s respiratory tract land on a recipient’s mucosal surface. Indirect droplet spread happens via direct physical interaction with of an infected person, or of the surfaces and fomites that have contacted with the infected person or their droplets.^13^ On the other hand, airborne transmission occurs when aerosolized particles are inhaled into the lower airway. Although it is often stated that droplets with a diameter (*d_p_*) > 5 μm can deposit directly on nearby surfaces while smaller aerosol particles (*d_p_* < 5 μm) remain in the air for significantly longer periods of time, the distance a particle travels is complex and dependent on many dynamic variables such as particle size, shape, charge, flow velocities, composition, density, air turbulence, temperature, and humidity.^14–16^

Patients with respiratory infections often generate large amounts of aerosols from coughing, sneezing, or heavy breathing.^17,18^ Some studies have also shown an association between aerosol-generating procedures and healthcare provider infection during the SARS-CoV-1 epidemic.^19,20^ Faced with likely risk of airborne transmission and lack of proper protection, many health care professionals have developed their own equipment and guidelines to protect themselves from the infection when working with patients.^21,22^ Existing devices such as the “intubations shield” (**Figure 1B**) are limited to a physical barrier of protection, as there are currently no strategies that provide protection from aerosol particles generated by a patient’s coughs and sneezes, and by high-risk procedures performed by healthcare providers.^21^ In the face of great urgency and severe risks, we have designed, constructed and tested the effectiveness an affordable, scalable Box for Aerosol and Droplet Guarding and Evacuation in Respiratory infection (BADGER) to contain both droplets and aerosol particles (**Figure 1C**).

**Figure 1:**
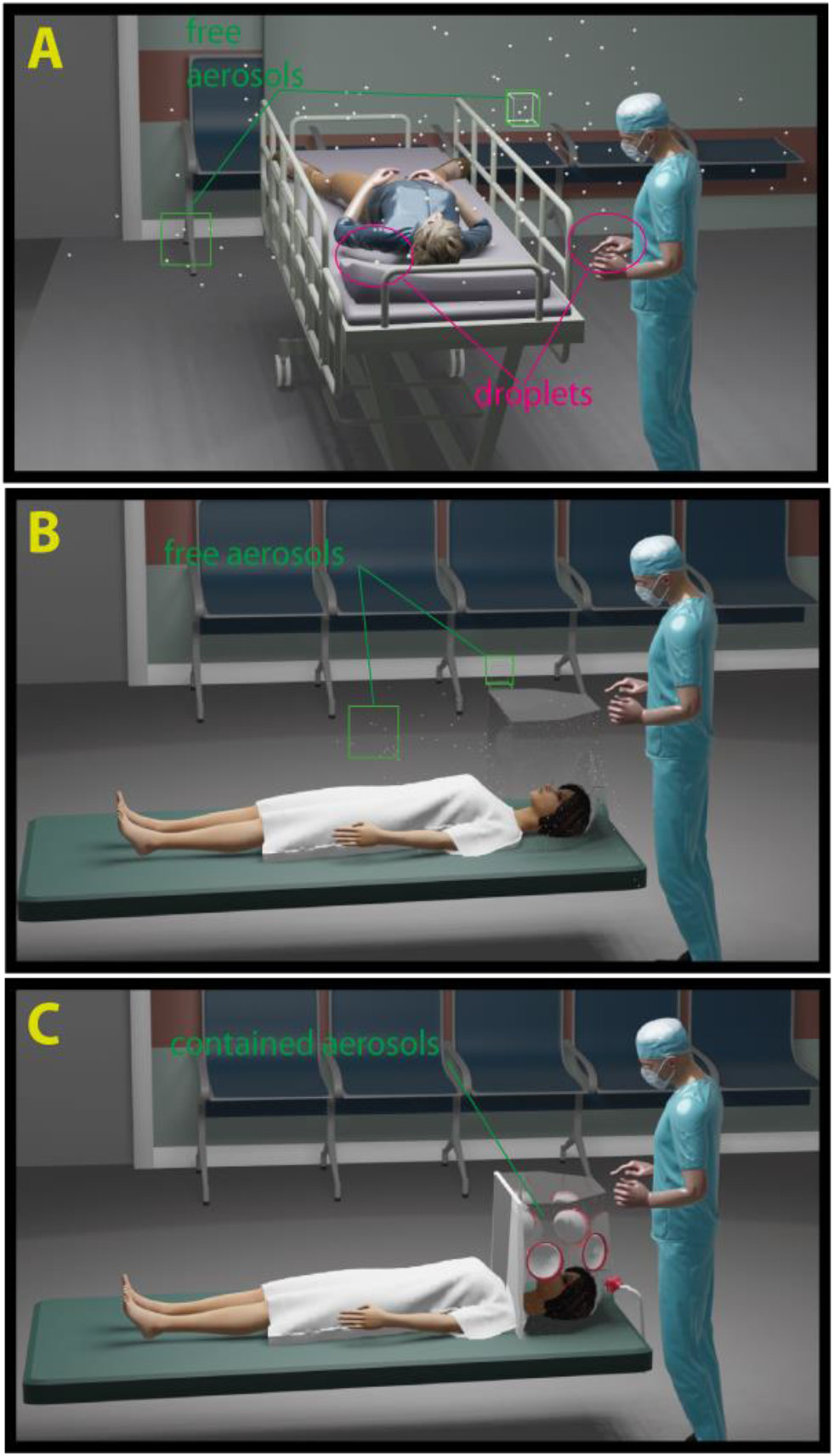
**(A)** Proposed routes of transmission of COVID-19. Spreading of respiratory infected disease as suspected in a COVID-19 patient by direct and indirect droplet contact (magenta ovals), and airborne via aerosol particles (green squares). (**B**) A regular intubation shield can stop the majority of large droplets. However, it is unable to contain aerosol particles (green squares) due to open design. (**C**) The BADGER with all accessories assembled and suction turned on, can act as both as a shield and a containment chamber, trapping aerosol particles inside until they are suctioned out.

## Methods

### Construction of the BADGER

To decrease risk of airborne transmission of SARS-CoV-2 to health care professionals and workers, we sought to build a device that could decrease exposure of providers to the patient’s droplets and aerosols.

We defined the following criteria for our device to meet the function, urgency and demand during the pandemic: 1) ability to stop direct droplet contact, especially from a patient’s coughs and sneezes, or during high risk procedures such as intubation of an airway and extubation; 2) ability to contain aerosol particles; 3) ease of manufacturing; and 4) affordability.

We studied the “aerosol box” developed by Dr. Hsien-yung Lai from Taiwan and its ability to shield providers from large flashing droplets.^21,23^ Although innovative and simple, this device lacks the ability to adequately contain aerosolized virus that could be present within smaller diameter aerosol particles due to its open air design (Figure 1B). In order to contain smaller diameter airborne particles, a negative pressure and/or rapid air exchange rate is needed, similar to the function of a negative-pressure room. To achieve our goal of keeping the device affordable and easy to manufacture, we used commonly available materials, clinical equipment and supplies. After three prototypes, we reached a production version. The BADGER is constructed of a clear reusable shell and disposable accessories and, when assembled, creates a closed chamber. The shell of the BADGER is a tapered pentagonal box that has a height of 50cm, a torso-opening of 60cm, and overall volume of 95.8 liters (L) (**Figure 2**). There are 4 hand-ports for health care providers and assistants, and two 22mm diameter openings for two viral high efficiency particulate air (HEPA) filters placement. The side panels are constructed by bending, or by cutting into separate panels (**Figure 2B**). The top panel is glued to the side panels using a hot melt adhesive (**Figure 2C**). To create a semi-sealed chamber, all the openings need to be closed off. For the hand-ports, we used the sleeve-ring method by assembling large disposable surgical gloves on 3D custom-made printed grommets (**Figure 2D**).

**Figure 2:**
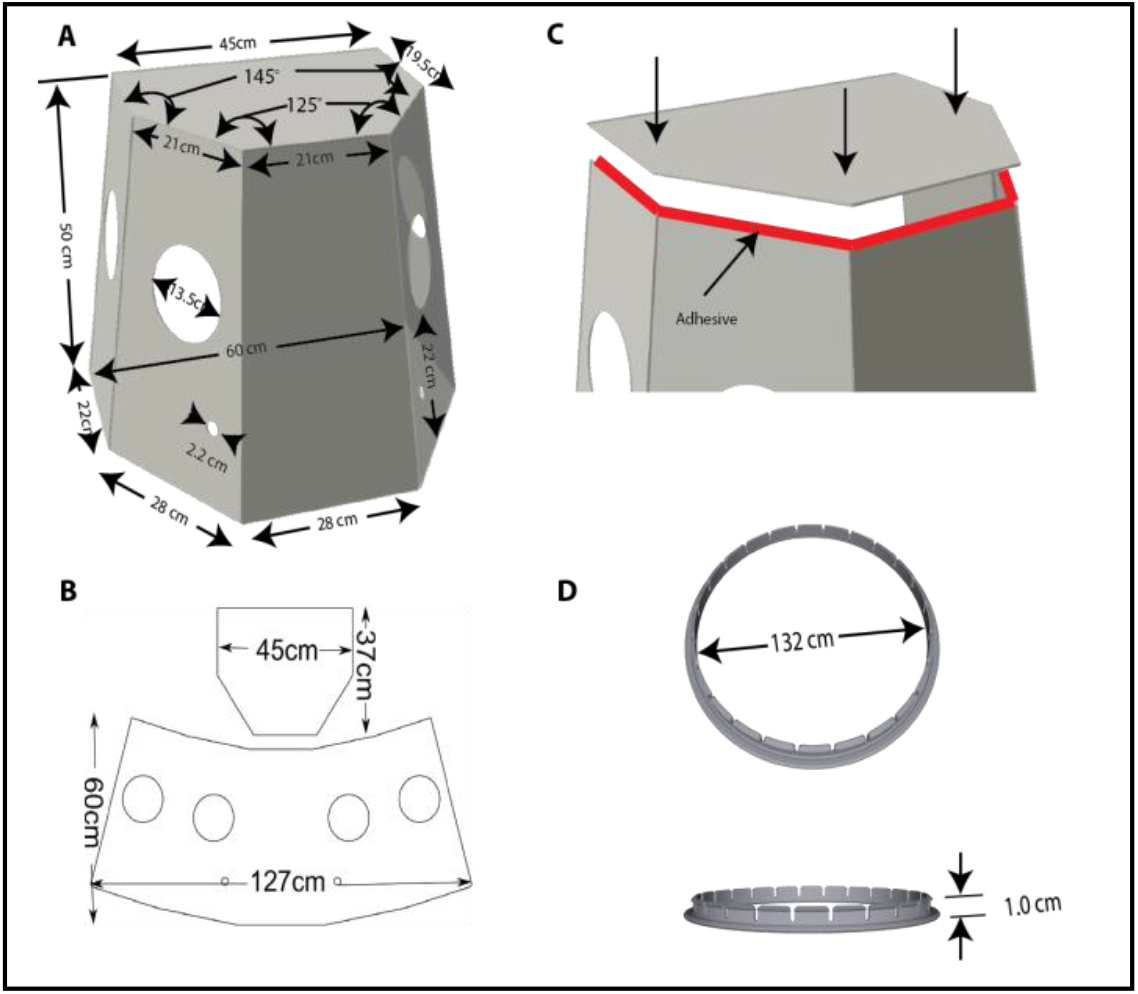
Configurations of the BADGER and grommet. (**A**) The BADGER has a tapered pentagonal shape with an overall height 50 cm and overall volume 95.8 L, four 135 mm diameter hand-ports and two 22 mm diameter outlets for filter placement. **(B)** The side panels can be constructed via bending or cutting into separate panels. (**C**) Top panel is glued to side panels using hot melt adhesive. **(D)** 3D-printed grommets, used to attach surgical gloves to the BADGER.

The glove fingers can be trimmed off at the palm to create a sleeve. While this compromises the seal at the hand-ports, it allows greater mobility, reach, and reduces wrong-handedness. The sleeve-rings are then inserted into the hand-ports of the BADGER (**Figure 3**). The large interior opening for the torso is sealed off from the outside environment using two overlapping plastic drapes. Large diameter tubing such as ventilator tubing is used to evacuate air from the box after passing through the HEPA filters.

**Figure 3:**
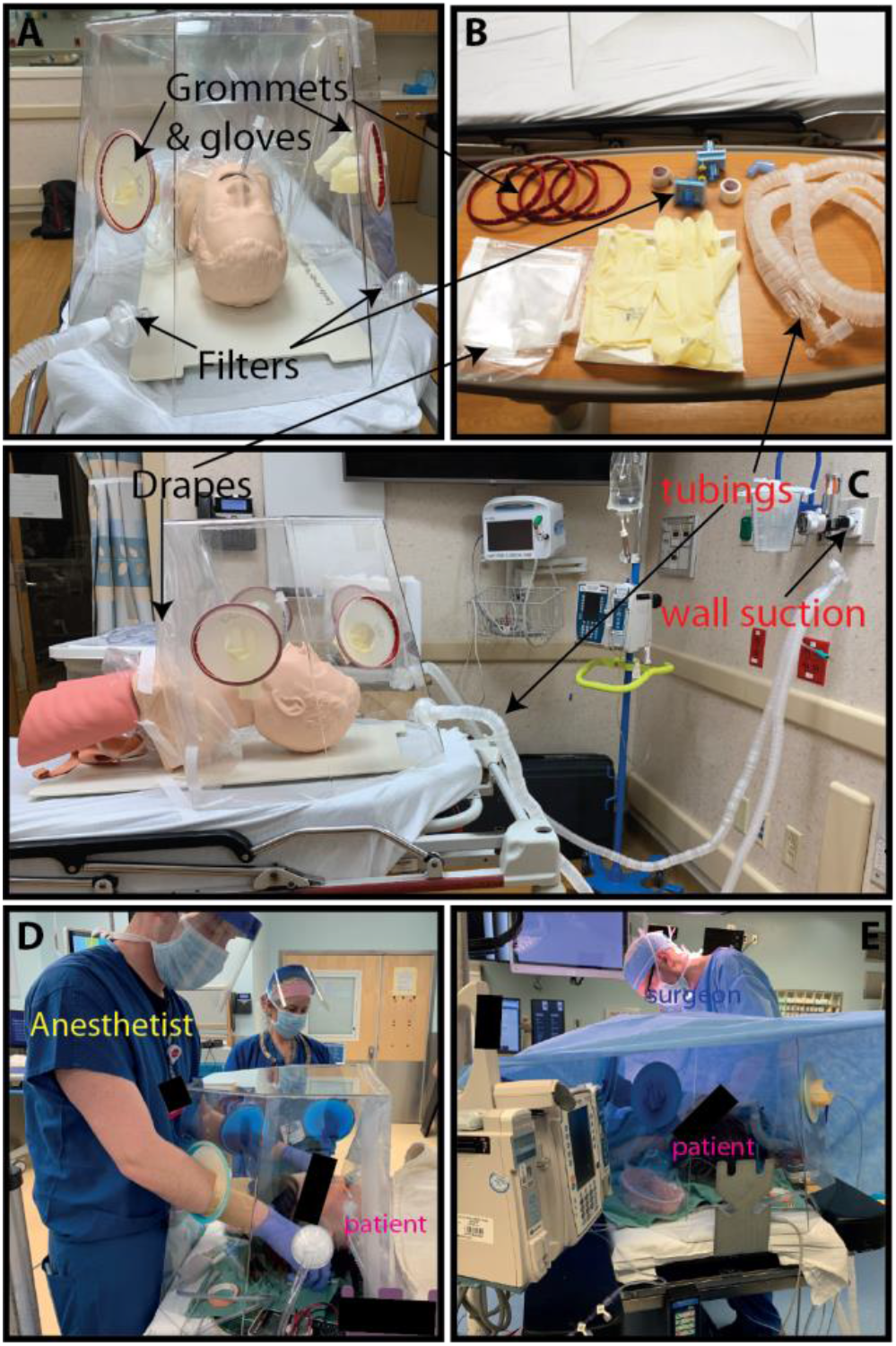
**(A-C)** Set up of the BADGER on a mannequin. The BADGER uses commonly available hospital supplies to create isolation and semi-sealed chamber. Accessories for each BADGER are: two viral HEPA filters, two 60 cm x 60 cm clear drapes, 5-10 m of 6 mm or 7 mm inner diameter tubing, one dual-limb breathing circuit tubing, 2 pairs of size 8.5 or 9 surgical gloves. Grommets and surgical gloves are assembled into sleeve-rings to seal the hand-ports. Drapes are taped to create overlapping opening for the torso/neck. (**D-E)** Clinical application of an earlier version of the BADGER on a patient for intubation (**D**) and intraoperative use (**E**).

### Outflow using wall suction

To model the BADGER after a negative pressure room, we designed the device to have a minimum of twelve air changes of exhaust per hour.^24^ To achieve this goal, we connected the BADGER to a standard hospital room’s in-wall vacuum suction source. In most modern hospitals, vacuum pressure is set to be at least 277 mmHg (36,930 Pa) which is 483 mmHg below atmospheric pressure at sea level.^25^ The latter number (atmospheric pressure - vacuum pressure) is a commonly used vacuum suction strength in a hospital or a clinical setting to quantify suction strength. We refer to this as suction pressure throughout the manuscript. However, we will use the former number (vacuum pressure) in calculation of flow rate. To maintain at least twelve air changes per hour (the BADGER’s volume = 95.8 L), the outflow rate must be at least 19.2 L per minute (L/min) or 9.6 L/min per filter outlet.

Air flow rates from a standard hospital room’s in-wall vacuum suction source was then measured using an in-line flow meter (TSI model 4043). The tests were carried out at room temperature (21°C) and atmospheric pressure (760 mmHg or 101.3 KPa). The flow meter was fitted to different types of commonly available suction tubing used in hospitals and clinics and a mixed setup: 1) non-sterile, non-conductive 7 mm inner diameter (D) tubing (Universal Argyle™ bubble tubing, Covidien); 2) sterile, non-conductive 6 mm D tubing (Cardinal Health, IL); 3) “Mixed setup” of a single 7 mm D tubing connected to a dual-limb breathing circuit tubing (Adult UltraFlex® Dual-Limb breathing circuit, King Systems, Noblesville, IN). We used 2 different lengths (5 and 10 m) as these would provide adequate length to connect the BADGER to an in-wall vacuum suction source. Flow rates were assessed with and without a viral HEPA filter: Ultipor25® (Pall, Port Washington, NY) or AG7178 Bacterial/Viral Filter (AG Industries, St. Louis, MI) attached to its upstream. We chose the “Mixed setup” to represent the current setup of the BADGER for our clinical use: 2 HEPA filters from the filter outlets connected to a dual-limb breathing circuit (1.8 m when stretch), which is in turn connected to a single 3.2 m × 7 mm D tubing. Measured volumetric air flow rates were plotted against the suction pressure settings that are commonly used in a clinical setting: low = 50 mmHg, medium = 100 mmHg, high = 150 mmHg, very high = 200 mmHg, and max suction = 700 mmHg.

### Modified Bitrex containment test

To investigate the BADGER’s ability to prevent aerosols from escaping and reaching surrounding personnel, we adapted a respirator fit test that is used to verify the seal of surgical respirators (e.g. N95 masks).27 Denatonium benzoate (Bitrex) is a non-toxic bitter chemical that can be detected by inhaling aerosolized droplets when breathing by mouth at concentration as low as 0.05ppm.28 Bitrex or saline control was nebulized inside of the BADGER in the standard operating configuration (**Figure 4A**). The test was carried out in a 5 m × 5 m well-ventilated non-negative pressure room. Volunteers, who have previously completed a respirator fit test and were able to detect the bitter taste, were asked by a blinded test administrator to detect any bitterness while breathing by mouth for 7 minutes while the sample was nebulized continuously into the BADGER. To compare the efficacy of the BADGER without seal and suction, nebulized Bitrex or saline were introduced while the drapes were lifted up and no suction was applied. The time to bitter taste detection was recorded.

### Negative pressure with smoke tests

To visually assess the BADGER’s ability to generate negative pressure, we conducted a series of smoke tests using ventilation smoke tubes (Grainger, Lake Forrest, IL) in a simulated hospital room at the University of Wisconsin-Madison Simulation Center (**Figure 4C-D**). The smoke test is a standardized test used to verify negative pressure in an enclosure (e.g., negative pressure room). White smoke generated by the smoke tube is easily visualized under standard room conditions. Smoke was introduced into the BADGER in the standard operating configuration. To simulate a non-invasive respiratory therapy, we placed a 10 L/min oxygen facemask on a high-fidelity mannequin, which was positioned in the BADGER with semi-sealed enclosure. We simulated coughs by quickly compressing the mannequin’s lungs (**Figure 4D**). The exterior of the BADGER was monitored by two observers to determine if there was smoke leakage.

**Figure 4:**
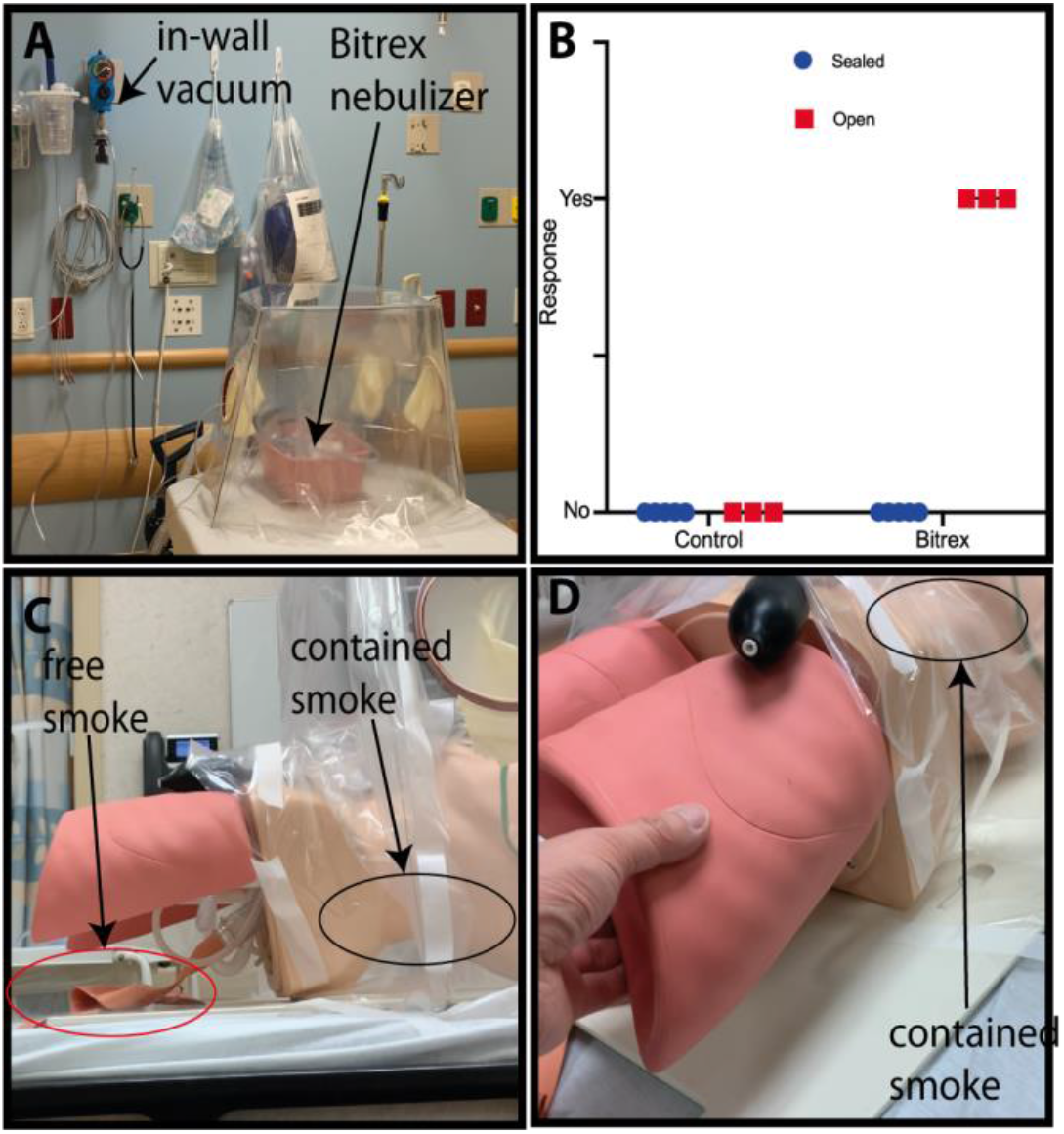
**(A**) Bitrex containment test setup with nebulized Bitrex or saline control inside the BADGER. Bitrex was delivered by oxygen nebulizer at 5 L/min. (**B)** Results of a double-blinded containment test. Zero out of 5 volunteers detected bitter taste when the BADGER was sealed, and suction applied at 100 mmHg to create outflow rate of 39 L/min. When the BADGER was unsealed and suction turned off, 3/3 in the Bitrex group and 0/3 in the saline group detected bitterness. (**C)** Smoke tests using ventilation smoke tubes (Grainger) while the mannequin was receiving 10 L/min of oxygen via facemask. Free smoke (red oval) was seen leaking from the bottom of the BADGER before vacuum suction was applied. (**D)** After suction was applied at 100 mmHg, coughs were simulated by compressing the mannequin’s lungs; all the smoke was contained (black oval).

### Sub-micrometer aerosol particle containment test

The ability of the BADGER for containing sub-micrometer aerosol particles was assessed by placing the BADGER into a secondary sealed enclosure. A controlled aerosol source atomizer (TSI model 3076) was placed inside the BAGER to simulate aerosol production. Aerosol particle concentrations are measured inside both the BADGER and the exterior enclosure. Aerosol particles from the output of the atomizer were dried to a relative humidity (RH) <10%. The total output aerosol concentration added to the BADGER was *ca* 1 × 10^6^ particles/cm^3^ at a flow rate of 2.6 L/min. Aerosols generated from the atomizer had a narrow size distribution from 10 to 300 nm, with a mean diameter of 65 nm.

The secondary sealed enclosure (0.45 m^3^) was purged continuously with particle free air at 16 L/min. Measurements of aerosol particles were made with a condensation particle counter (CPC, TSI model 3787) and a scanning electrical mobility spectrometer (SEMS, Brechtel model 2100). The CPC measures total aerosol particle concentration (particles/cm^3^) for particles from 5 nm to >3 μm. The CPC sample flow rate was 1.5 L/min. The SEMS measures aerosol concentration and size distribution for particles from 10 nm to 1.4 μm. The SEMS sample flow rate was 0.31 L/min and supplemented with an additional pump pull of 1.2 L/min to create a total flow of 1.5 L/min matching the CPC. All aerosol sampling lines were composed of stainless steel and black conductive tubing. Sampling lines to both the SEMS and CPC were approximately 2.5 m long. Sampling lines for the CPC and SEMS were plumbed with valves to enable each instrument to alternate sampling from either the BADGER or the outer enclosure. An aerosol source line from the atomizer was run into the center of the BADGER to enable introduction of a stable aerosol particle source. The default configuration of the BADGER in these tests included two AG7178 Bacterial/Viral filters connected in parallel to the vacuum source with a controllable flow rate. All hand-ports were sealed. The plastic drapes were installed in standard operation configuration.

The standard experimental procedure followed the following sequence: 1) Purge the outer enclosure until a particle concentration of <50 particles/cm^3^ was measured in both the BADGER and the outer enclosure. 2) Start the 2.6 L/min aerosol particle source flow into the BADGER and sample aerosols in both BADGER and outer enclosure until a stable signal is reached. 3) Shut off the aerosol source flow and record the particle decay in the BADGER and outer enclosure. 4) Repeat at different filter pump rates or box configurations. This procedure enables determination of the aerosol concentration ratio in the BADGER (*N_inner_*) and the outer enclosure (*N_outer_*), which was used to calculate the fraction of particles that were contained in the BADGER.

### Filter Efficiency Testing

The filter efficiency of aerosol particles was determined for several standard HEPA filter types as a function of flow rate through the filter. Aerosol particle concentrations and size distributions were measured with the SEMS. The inlet of the SEMS was connected to a dry scroll pump to vary sample flow rates through the filter from 1.5 to 25 L/min. The flow rate was measured with an in-line flow meter (TSI model 4143) downstream of the SEMS sampling point. Aerosol concentration and size distribution were first determined on laboratory room air prior to testing of each filter. Laboratory air particle concentrations were typically from 1000 − 2000 particles/cm^3^, with a mean diameter of 220 nm. Filters were connected to the SEMS inlet with black conductive tubing. Room air was sampled through the filter at multiple flow rates in order to determine the flow rate when particle breakthrough occurred for the filter. Filter efficiencies were determined by dividing aerosol concentration through the filter by the room air aerosol concentration, expressed as a percentage. The three standard filter types tested were: Ultipor® 25, Pall BB22-15, and AG7178 Bacterial/Viral filters. The AG7178 filter was also tested as a function of filter number (1 or 2) and filter arrangement (in series or in parallel).

The need for approval and consent from the Institutional Research Board of the University of Wisconsin-Madison were waived because the study was considered a quality improvement project.

## Results

### Construction of the BADGER

A production version of the BADGER can be constructed from a variety of transparent or semi-transparent materials. Clear materials that are appropriate are acrylic (PMMA), polycarbonate (Lexan), PETG, or vinyl. Each material has advantages and disadvantages, as summarized in **Table 1**.

**Table 1:** Commonly available materials to construct the outer shell of the BADGER and their advantages and disadvantage in manufacturing and clinical use.

|  | Polycarbonate | Acrylic | PETG | Coroplast | Vinyl sheet |
| --- | --- | --- | --- | --- | --- |
| **Transparency** | Excellent | Excellent | OK | Poor | OK |
| **Bending** | Cold bending<br>Heat bending | No cold bending<br>Heat bending | Folds easily | Corrugated, must be<br>score/routed for bends<br>against corrugation | Folds easily |
| **Fabrication** | Router | CO2 laser<br>Router | Tangential blade<br>Router<br>Scissors | Tangential blade<br>Router<br>Scissors | Tangential blade<br>Router<br>Scissors |
| **Durability** | Durable<br>Prone to scratching | Crack prone/<br>shatters<br>Scratch resistant | Durable<br>Scratches easily | Durable<br>Scratches easily | Soft<br>Scratches easily |
| **Bonding** | Hot melt glue<br>Silicone caulk<br>Methylene chloride | Hot melt glue<br>Silicone caulk<br>Solvent welding | Double sided tape<br>Tape<br>Sewing | Double sided tape<br>Tape | Double sided tape<br>Tape<br>Sewing |
| **Cleaning** | No Abrasives | No alcohols or<br>solvents | No Abrasives | No Abrasives | No Abrasives |

Although the shell of the BADGER should be built by facilities with appropriate tools such as a computer numerical control (CNC) router, laser cutter, or die/blade cut, it can also be built using a box cutter and a metal straightedge. Our systems were constructed from CNC routed polycarbonate. Polycarbonate is an extremely durable material. Although it is slightly softer and more prone to scratches than acrylic, it is more chemically stable and easier to clean. Generally, polycarbonate is slightly more costly than acrylic. However, the cost of raw materials for this shell is still very affordable (<$100 material cost/unit). The tapered shape of the BADGER also allows it to be stacked, making the units easier to store and ship. Manufacturers can easily scale up the production if demand is high. Technical support for manufacturing, open-source design drawings, and lists of materials are available at the University of Wisconsin-Madison’s MakerSpace.^29^

### Outflow rate from a hospital room’s in-wall vacuum suction

Using commonly available suction tubing connected to a standard hospital room’s in-wall vacuum suction source, we obtained a minimum outflow rate of 16.5 L/min when using 10 m length × 6 mm D tube connected to low suction (50 mmHg) without filter and 14.5 L/min with an AG7178 Bacterial/Viral filter. Both AG7178 and Ultipor® 25 performed similarly in impeding air flow. The outflow rate increased to 32 L/min at high suction pressure (200 mmHg) with filter and topped out at 41 L/min at maximum suction pressure (700 mmHg). As expected, outflow rate increased as tube length decreased or increased tubing D. However, the relationship between tube length and suction pressure to outflow rate are not linear (**Figure 5A**). In the mixed setup, we obtained a total outflow rate of 28 L/min at 50 mmHg suction pressure and maximum outflow rate of 59.7 L/min at 700 mmHg.

**Figure 5:**
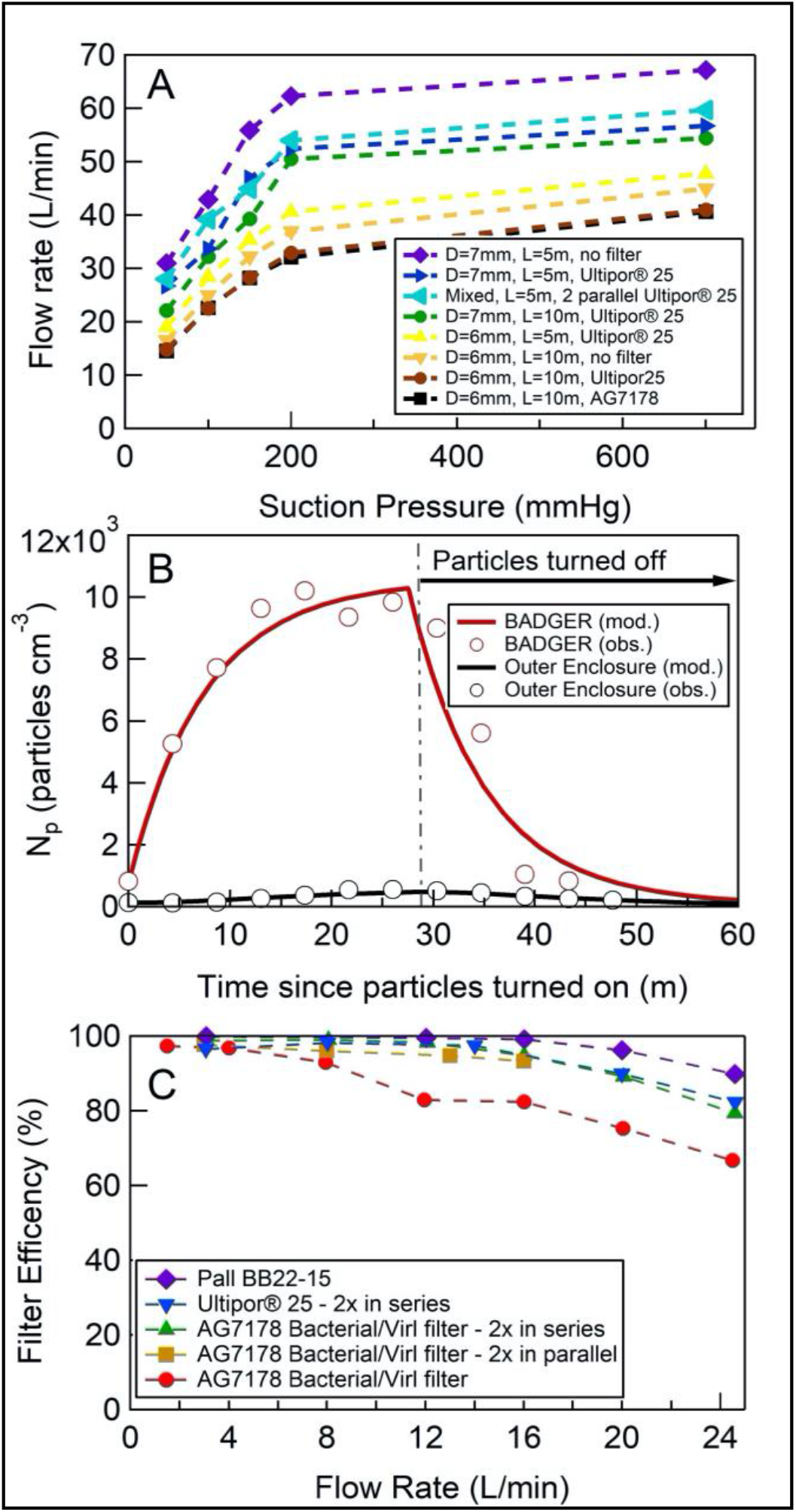
**A)** Air flow rates from a hospital room’s in-wall vacuum suction source at various suction pressures, tubing inner diameters (D), tubing length (L), and viral filters (Ultipor25® or AG7178). Mixed setup: 2 Ultipor 25® filters connected in parallel to 1.8m of a dual-limb breathing circuit tubing (Adult UltraFlex® Dual-Limb breathing circuit) and 3.2 m of 7 mm inner diameter tubing. **B)** Particle containment result at outflow rate of 13 L/min. Sub-micrometer particle concentrations were measured in both the BADGER (red circles) and the exterior enclosure (black circles). For the well-sealed BADGER, greater than 90% of sub-micrometer particles generated in the BADGER were contained. **C)** Measured aerosol particle filtration efficiency for the selection of filter substrates and combination of filters used in this study. The filtration efficiency test was conducted using aerosol particles present in the room, where the mean particle diameter was 220 nm.

### The BADGER contains Bitrex

When sealed off and suction applied at medium strength (100 mmHg) using the mixed setup to achieve approximately 39 L/min of outflow, the BADGER prevents nebulized Bitrex from leaking into the room, thus preventing 100% of blinded volunteers (n=5) from detecting its bitter taste. When the drapes were lifted off and suction was turned off, all blinded volunteers were able to detect the taste in the Bitrex group (n=3) in an average of 39 seconds, while none of the volunteers in the saline group detected bitter taste (n=3) (**Figure 4B**).

### Negative pressure with smoke tests

A series of smoke tests were conducted to visually assess the BADGER’s effectiveness in containing airborne particles by its negative pressure. In the first scenario, the mannequin was receiving 10 L/min of oxygen via a facemask. As smoke was introduced into the BADGER in the standard operating configuration without suction applied to the BADGER, there was visual leakage of smoke from under the drapes and from the hand-ports (**Figure 4C**). When suction was applied at medium strength (100 mmHg) to create an outflow rate of approximately 39 L/min, smoke was well contained inside the BADGER. When simulated coughs were performed by suddenly compressing the mannequin’s lungs, the BADGER was able to contain smoke inside its chamber (**Figure 4D**). Five minutes after cessation of smoke generation, the BADGER was able to clear the visible smoke inside.

### Sub-micrometer aerosol particle containment test

Results from a typical particle containment test (outflow rate of 13 L/min with filters) are shown in **Figure 5B**, where sub-micrometer particle concentrations were measured in both the BADGER (red circles) and the exterior enclosure (black circles). Particle concentrations in the BADGER approach steady-state following approximately 15 minutes, peaking at 1 × 10^4^ particles/cm^3^, while particle concentrations in the outer enclosure peak at < 500 particles/cm^3^. To determine the fraction of particles contained by the BADGER, we model the time dependent particle concentrations in each box, constrained by the measured flow rates into and out of each enclosure and the time dependent change in particle concentrations during a typical experiment. We utilize both the steady-state concentrations and the rise and fall times in both enclosures to determine the leak rate of particles from the BADGER to the outer enclosure and the particle wall loss rate in the BADGER. Model results are shown with solid lines in **Figure 5B**. Model-measurement agreement is achieved with a particle wall loss rate in the BADGER (*kwaii*) of 1 × 10^-4 s-1^ and a leak rate of 1.5 L/min from the BADGER to the outer enclosure. This results in containment of greater than 90% of sub-micrometer particles generated in the BADGER for the select operating conditions tested here. Particle containment was equally effective over the range of particle sizes generated by the particle source (10 to 300 nm).

### Filter efficiency testing

The filter efficiency of aerosol particles was determined for several standard particle filter types as a function of air flow rate through the filter. The three standard filter types were tested: 1) Ultipor® 25, 2) Pall BB22-15, and 3) AG7178 Bacterial/Viral filters. AG7178 filter was also tested as a function of filter number (1 or 2) and filter arrangement (in-series or in-parallel). Full results of the filter efficiency testing are shown in Figure 5C. Filter efficiency was greater than 90% at flow rates less than 18 L/min for all filter types except for the single AG7178 filter.

## Discussion

In response to the pandemic, unanticipated PPE shortages and lack of understanding about the transmission and infectivity of the SARS-CoV-2 virus has created the need for new tools to reduce risk of healthcare provider exposure based on best available information. Moreover, there is evidence that current PPE might not provide adequate protection against infectious airborne particles.^30^ Given the rapidly evolving situation, proof-of-principle devices have emerged to provide organizations with possible designs for combating the imminent challenges they are facing. Several physical barrier designs to protect healthcare providers during aerosol-generating activities have been developed. However, these devices largely limit protection to direct contact with larger droplets and lack proven efficacy.^21^ The use of negative-pressure isolation rooms is important to ensure infectious agents do not spread through the surrounding environment. However, many hospitals are not adequately equipped to handle the surge in patients during a pandemic.^31^

We designed, constructed, and tested a portable, affordable and easy to manufacture device (BADGER) which can contain aerosol particles produced by patients with respiratory infection such as COVID-19. The BADGER utilizes negative pressure to prevent dispersion of aerosolized virus throughout the room which could expose healthcare providers several hours later.^32^ We have performed both qualitative and quantitative tests; all demonstrated its effectiveness in containing aerosols particles. In this study, the aerosol dispersion test was conducted at lower outflow rate (13 L/min) compared with the recommended minimum outflow rate of 19.2 L/min. Therefore, we expect that at higher outflow rates and for larger aerosol particles, which are more representative of patient-generated aerosol particle diameters, the containment ability will improve even more significantly.

The results on the filter efficiency test indicated that filters, especially the AG7178 Bacterial/Viral filters, have reduced efficiency at flow rate more than 18 L/min. Multiple filters should be used in parallel to achieve high flow rate while minimizing aerosol particle breakthrough on the filters. Although viral HEPA filters reduce flow rate by approximately 15% in our testing, they are essential components if the exhausted air is to be reintroduced into the surrounding environment. Moreover, exhausted air from hospital in-wall vacuum is released without being filtered for virus or bacteria (personal communication). Therefore, we recommend always using viral HEPA filters for exhaust air from the BADGER.

The BADGER is designed to contain and evacuate aerosol particles at minimal outflow rate of twelve air changes of exhaust per hour. This will be particularly useful during aerosol-generating activities and procedures in compliant patients who can lie in the BADGER to reduce the risk of aerosol dispersion and transmission of respiratory infective particles to healthcare professionals and workers. As a result, institutional approval for clinical use has been granted by the University of Wisconsin Health during this pandemic. We have used the BADGER in the perioperative period with the consent of patients and their families (**Figure 3D-E**). The box was appropriately positioned prior to induction of anesthesia, and direct laryngoscopy was performed on all cases. There were no complications and endotracheal positioning and intubation were confirmed by capnography. The BADGER remained in place for the duration of all the cases. At the end of the operations, the patients were extubated inside the BADGER and had uneventful post-operative courses. As there is more evidence of aerosolized SARS-CoV-2 from innocuous activities such as sneezing and coughing, and current standard PPE might not fully prevent exposure for healthcare providers, the BADGER might be an additional layer of protection to mitigate airborne transmission of COVID-19.^11,30,32^

There are several limitations of this study and of the BADGER. It is foreseeable that in situations involving a difficult or obstructed airway, the device could limit access to the patient. In this scenario, the device could be easily lifted up and removed to restore complete exposure. The clear polycarbonate material allows for easy visualization of the patient’s airway by the anesthesia provider, and intubation could be performed by either direct laryngoscopy or videolaryngoscopy. However, the BADGER can still make procedures more difficult in critical situations. We recommend standardized, simulated training for those who will use this device before implementing in a clinical setting. We found that most users became very comfortable after 2-3 hand-on practices.

The optimal effectiveness of the box to limit aerosol dispersion is dependent on operating conditions including the flow rate of the vacuum suction and negative pressure generated. We have not tested the BADGER using portable vacuum pumps. Since portable vacuum pumps might not have a pressure regulator, thus limiting our ability to control outflow rate and prevent filter damage, this might limit the use of the BADGER to the settings where an in-wall facility-generated vacuum suction course is available. We have not extensively tested the BADGER using all tubing sizes and filter configurations at different vacuum levels. Although ideal outflow rate *Q* for laminar, isothermal, non-compressible gas can be estimated based on the Hagen-Poiseuille equation: 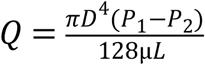 where *D* is the inner diameter of the tube, *P*_1_ is atmospheric pressure, *P^2^* is vacuum pressure, *L* is total length of tubing and μ is dynamic viscosity of air at room temperature and atmospheric pressure (1.81 × 10^-5^ Pa.s), we expected and confirmed that actual outflow rates are much lower. This is due to very high Reynolds number for high air flow rate indicating non-laminar flow and increases in Mach number at higher pressures due to compressible air.^26,33,34^ Flow rate saturates for each tubing configuration at approximately 200 mmHg vacuum (Figure 5A) indicating higher order compressibility and turbulent flow effects, which are not modeled by Hagen-Poiseuille, influence the response. In our experience, the mixed setup (2 HEPA filters, 1.8 m of ventilator tubing and 3.2 m of 7 mm diameter suction tubing) would provide good balance between outflow rate, filter efficiency and easy of setup.

While the BADGER provides a relative negative pressure analogous to a negative pressure room, the negative pressure is minimal, and these results are preliminary and purely in simulated environments. It is possible that the fraction of sub-micrometer particles contained during aerosol-generating activities and procedures (e.g., sneezing, intubation) may decrease as there is a sudden pressure increase inside the BADGER or as it becomes less well-sealed. Further testing must be performed to determine aerosol containment and clearance in more clinically relevant scenarios, both simulated and real clinical settings. In the meantime, healthcare providers should maintain appropriate respiratory PPE usage according to their institutional protocols. Furthermore, it is possible that CO^2^ levels in the BADGER could rise when a patient is inside. Since data on CO^2^ emission rate from a person inside the BADGER are not available, we recommend maintaining adequate outflow rate and frequent monitoring of the patient’s condition to avoid possible negative outcomes.

## Data Availability

Data will be made available upon request.

## Acknowledgements

University of Wisconsin-Madison Engineering Support for Covid-19 for engineering support.

American Family Children’s Hospital Operating Room for supplies and equipment support.

Russel Ward, Tuan Nguyen, Cari Myers, Elizabeth Yu, Scott Springman, Department of Anesthesia and Joshua Ross, Department of Emergency Medicine for clinical testing and feedbacks.

Shannon DiMarco, The Simulation Center at University of Wisconsin School of Medicine and Public Health for simulation support.

Greg Nellis, Department of Mechanical Engineering for help with air flow testing.

Tony McGrath, Biological Safety Cabinet Program of University of Wisconsin-Madison and Ben Eithun of University of Wisconsin Health for help with smoke test.

Christopher D. Cappa, Department of Civil and Environmental Engineering, University of California – Davis for helpful discussion on particle transmission and loan of the SEMS instrument.

William Murphy and Dana Maya for manuscript discussion and editing support.

Author Contribution:

Concept and Design: HDL, THB

Acquisition, analysis, or interpretation of data: all authors

Drafting of the manuscript: HDL, GAN, JW, KJ, KH, ELO, THB

Critical revision of the manuscript: HDL, KJ, CM, AW, THB

Administrative, technical, or material support: HDL, GAN, JR, CM, KJ, ELO, THB

Supervision: HDL

